# A hybrid approach to predict COVID-19 cases using neural networks and inverse problem

**DOI:** 10.1101/2022.05.17.22275205

**Authors:** Subhendu Paul, Emmanuel Lorin

**Affiliations:** School of Mathematics and Statistics, Carleton University, Ottawa, K1S 5B6, Canada; Centre de Recherches Mathématiques, Université de Montréal, Montréal, H3T 1J4, Canada

## Abstract

We derive a novel hybrid approach, a combination of neural networks and inverse problem, in order to forecast COVID-19 cases, and more generally any infectious disease. For this purpose, we extract a second order nonlinear differential equation for the total confirmed cases from a SIR-like model. That differential equation is the key factor of the present study. The neural network and inverse problems are used to compute the trial functions for total cases and the model parameters, respectively. The number of suspected and infected individuals can be found using the trial function of total confirmed cases. We divide the time domain into two parts, training interval (first 365/395 days) and test interval (first 366 to 395/ 396 to 450 days), and train the neural networks on the preassigned training zones. To examine the efficiency and effectiveness, we apply the proposed method to Canada, and use the Canadian publicly available database to estimate the parameters of the trial function involved with total cases. The trial functions of model parameters show that the basic reproduction number was closed to unity over a wide range, the first from 100 to 365 days of the current pandemic in Canada. The proposed prediction models, based on influence of previous time and social economic policy, show excellent agreement with the data. The test results revel that the single path prediction can forecast a period of 30 days, and forecasting using previous social and economical situation can forecast a range of 55 days.

## Introduction

The novel coronavirus disease 2019 (COVID-19), the current pandemic, caused by the deadly virus SARS-CoV-2 continues to pose critical and urgent threats to global health. The outbreak reported first time in early December 2019 in the Hubei province of the People’s Republic of China has spread worldwide^1^. As of 26th October 2021, the overall number of patients confirmed to have the disease has exceeded 245,121,000 in more than 220 countries, although the number of people infected is probably much higher, and more than 4.9 million people have died from COVID-19^2^. This pandemic continues to challenge healthcare systems worldwide in many aspects, including the demands for hospital beds and critical shortages in medical equipment. Thus, the capacity for immediate clinical decisions and effective usage of health-care resources is crucial. In this circumstance to handle the situation, generated by COVID-19, it is essential to derive a reliable and long-time prediction.

There are numerous analytical and computational studies based on mathematical models, involving Ordinary Differential Equations (ODE)^3–13^ as well as Delay Differential Equations (DDE)^14–21^, to calculate the basic reproduction number *R*_0_ and understand the underlying dynamics of the epidemic. In addition to those mathematical approaches, there are several statistical studies^22–30^, based on various samples of patients such as severe, non-severe, ICU, non-ICU, large size, small size, meta-analysis, estimated the features of the current pandemic. Furthermore, researchers use neural networks^31,32^and inverse problem technique^33–36^ aiming to provide insight and understanding the trends of the current pandemic COVID-19. Evaluating coefficients and other parameters in a differential equation using observed data is the key of inverse problems. However, inverse problems are challenging to solve because as they do not provide unique solutions, and they require prior information. For illustration, suppose we know that the time dependent parameter *α*(*t*) is positive definite, so we can ignore all the negative solutions obtained by solving the inverse problem. This challenge can be treated as the only limitation of the present study, developing the prediction models for COVID-19 cases. Prediction models^37–39^ that combine several features to estimate the risk of infection have been developed, in the hope of assisting medical staff worldwide in trigging patients, especially in the context of limited health-care resources.

The proposed hybrid approach is an amalgamation of several stages: epidemic model, a second order nonlinear equation for the total cases, a trial function for total cases using Neural Network (NN), trial functions for the model parameters using NN plus inverse problem technique, and finally prediction model employing Taylor series.

Firstly, we start with a compartment based modified SIR model with five partitions; these are Susceptible, Infected, Confirmed cases, Recovered and Deaths. Total cases *T* is a sum of active cases, recovered individuals and death tolls. From the first order nonlinear model equations, we generate a second order nonlinear differential equation for *T* involving model parameters. As a consequence, instead of dealing with the system of model equations, we merely focus on that second order equation.

Secondly, we derive a trial function for *T* using a single layer NN, and the parameters of the NN are estimated using publicly available data^40^. The trial function transforms the discrete data into a continuous curve. From the trial function of *T* we can analytically evaluate the first and second order derivatives of *T*.

Thirdly, we develop the trial functions of the model parameters involved in the second order nonlinear equation using NN and inverse problem technique. We train the error function to obtain the estimated values of the parameters connected with the single layer NN. The trial function of the model parameters help to calculate the time dependent basic reproduction number *R*_0_(*t*). Time dependent model parameters correspond to the social economic condition such as lockdown/reopening. Suppose that at a particular time *t*_1_, the values of the model parameters *β* (*t*_1_) and *δ* (*t*_1_), involved in the second order nonlinear equation, correspond to a particular social economic condition. Now, if that particular social economic condition occurs in the future, we can consider the values of the model parameters to be *β* (*t*_1_) and *δ* (*t*_1_). This assists us to develop a prediction model.

Finally, we develop the prediction models using Taylor series; there are three terms in the prediction model, previous total cases, influence of the preceding time and effect of social economic policy. According to the choice of prior data/situation, we obtain two types of prediction models. The proposed forecasting model can capture the social economic policy and can use it to forecast the total cases.

## Results

The outcomes of the NN (Figure 1), inverse problem (Figures 2 and 3) and the prediction model based on Taylor series (Figures 4 and 5) are reported in this section.

**Figure 1.**
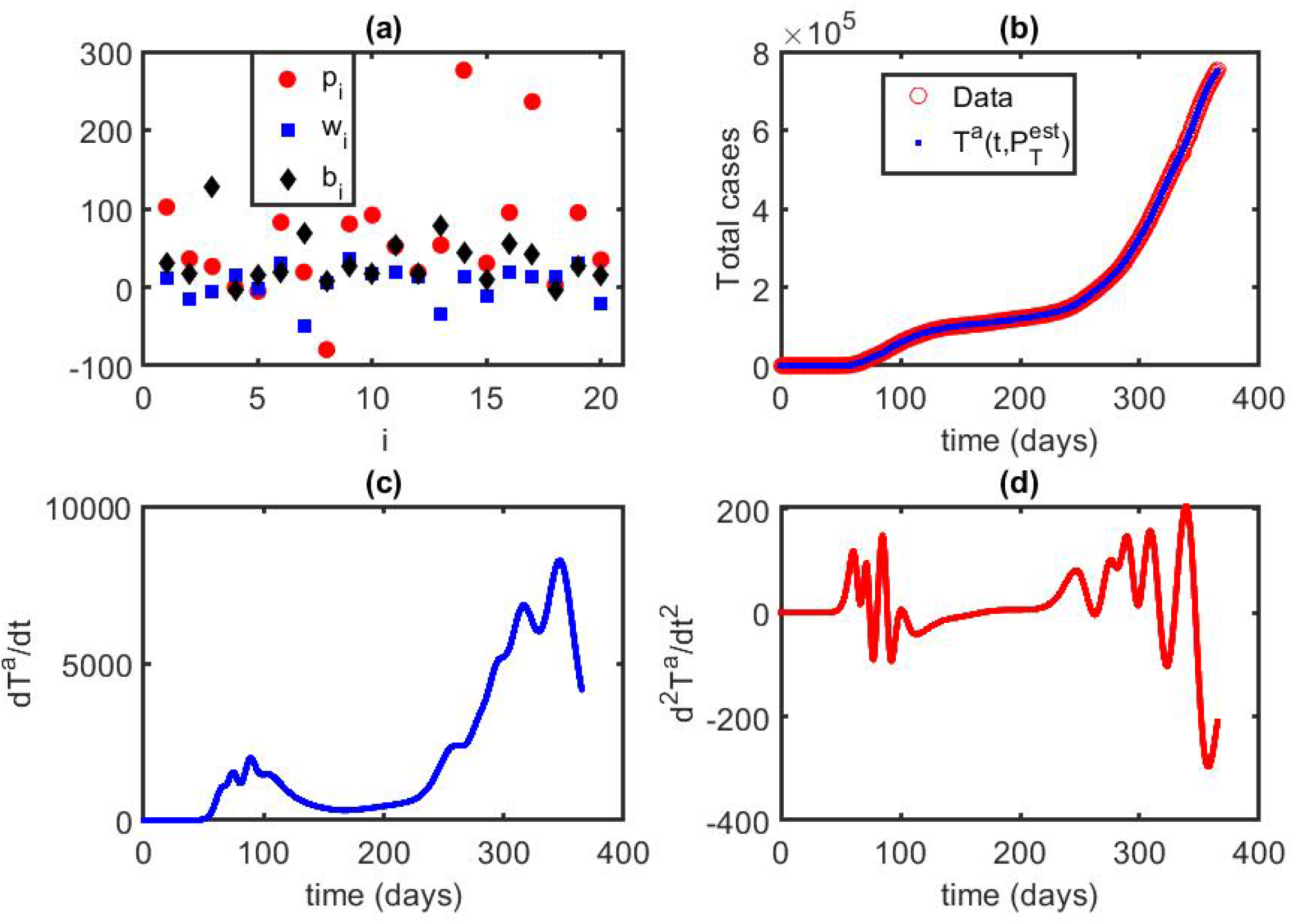
Total number of confirmed coronavirus cases: **(a)** Estimated values 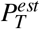of the NN parameters *P*_*T*_, involved in the trial function of total cases *T*^*a*^(*t*). The number of hidden unit is 20 so *i* = 1, 2, …, 20. **(b)** Trial function of total cases 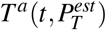 compared to the available data^40^. Blue dots indicate the results obtained from the NN, and red circles are the publicly available data^40^. **(c)** First order derivatives of the trial function 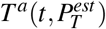. **(d)** Second order derivatives of the trial function 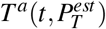.

**Figure 2.**
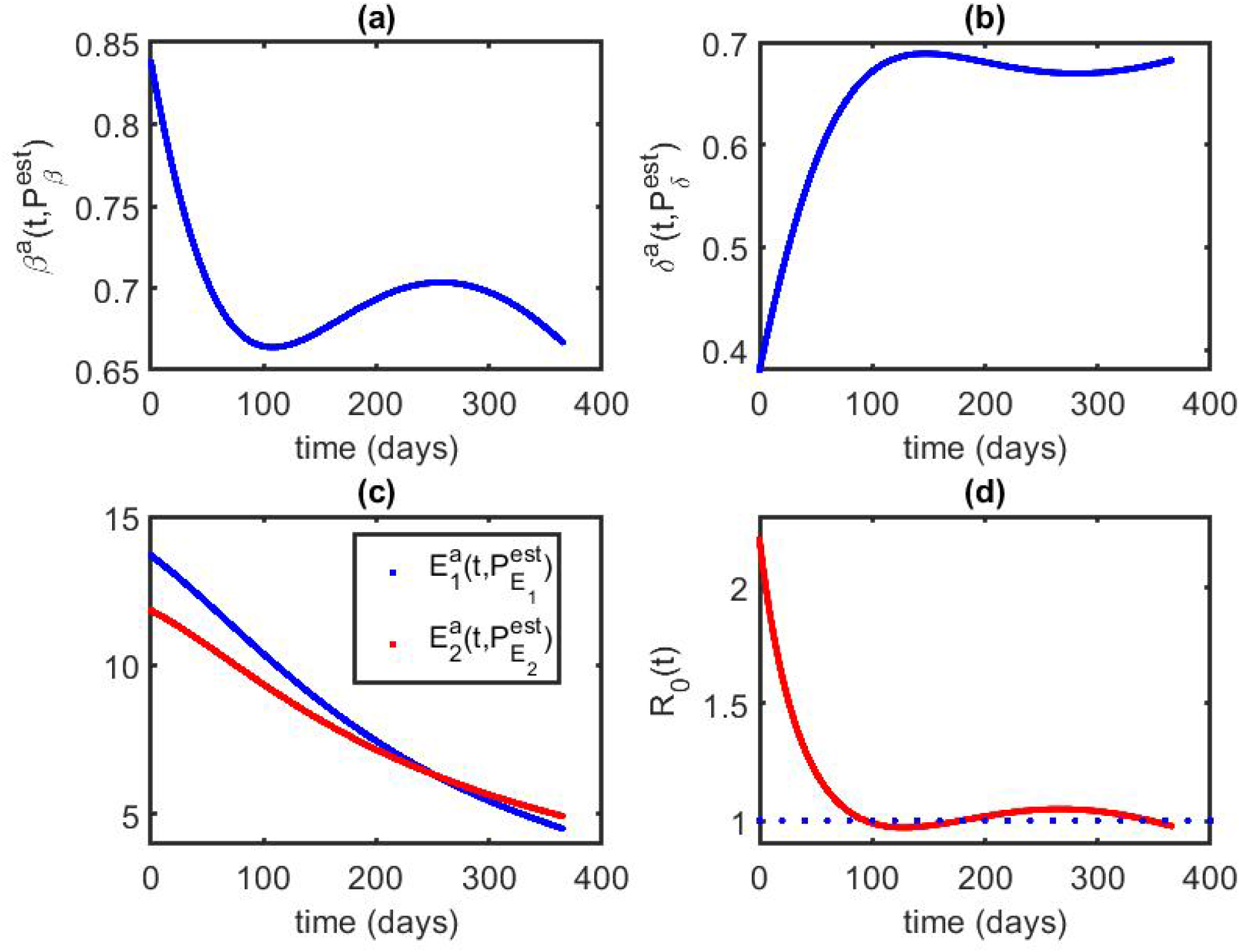
Outcomes of NNs and Inverse problem: **(a)** Trial function 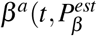 of the model parameter 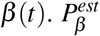 is the estimated value of the NN parameters *P*_*β*_. **(b)** Trial function 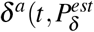 of the model parameter 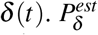 is the estimated value of the NN parameters *P*. **(c)** Trial functions of the error corrections *E*_*I*_ and *E*_*I*_. 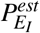 and 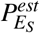 are the estimated values of the NN parameters 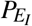 and 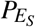 involved with the trial functions of *E*_*I*_ and *E*_*S*_, respectively. **(d)** Time dependent basic reproduction number *R*_0_(*t*) for the first 365 days of the current pandemic in Canada.

**Figure 3.**
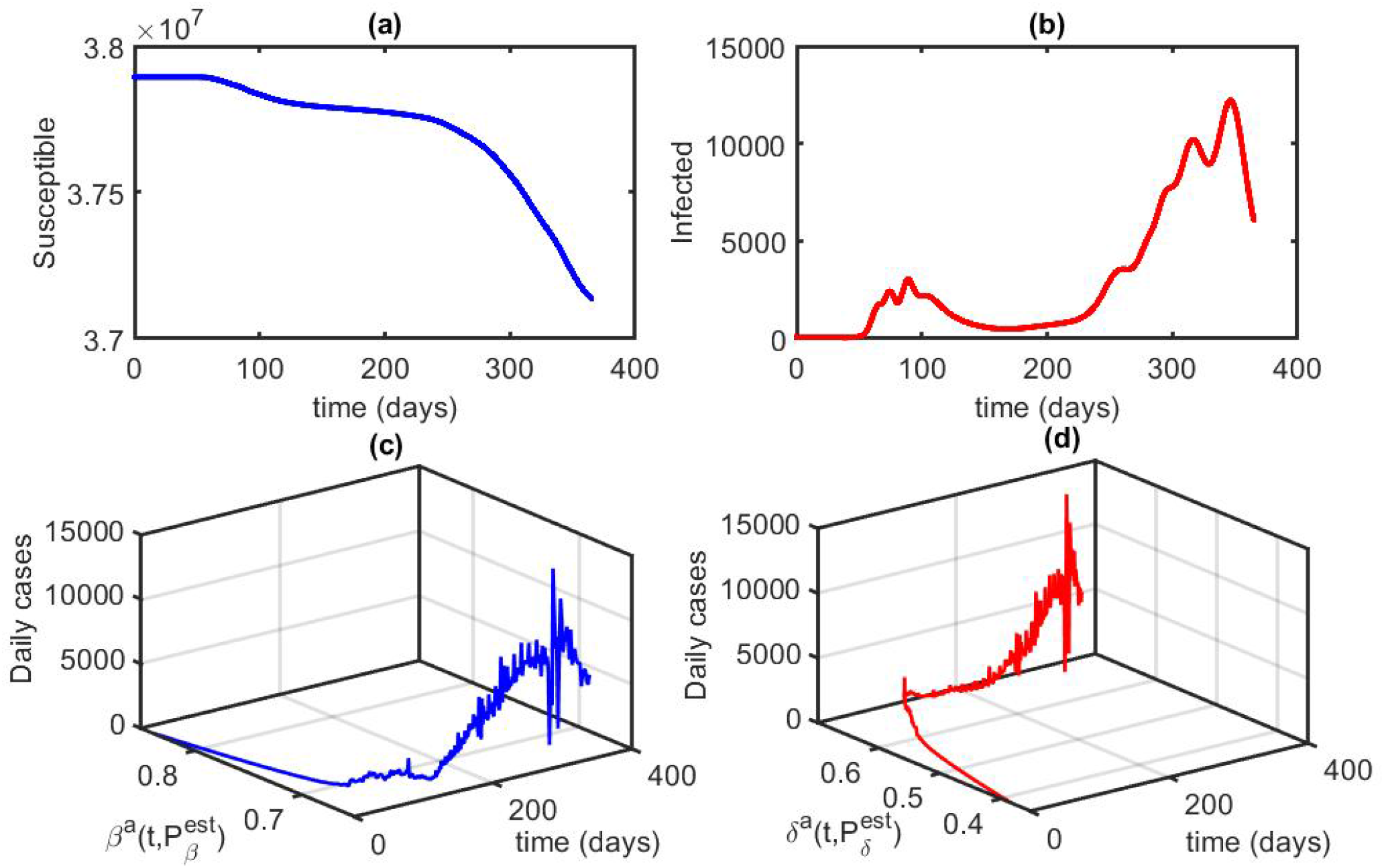
Outcomes of NNs and Inverse problem: **(a)** Number of susceptible individuals obtained from the publicly available data^40^ of the total confirmed cases. **(b)** Number of infected individuals obtained from the publicly available data^40^ of the total confirmed cases. **(c)** The trajectory (daily cases, 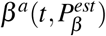). **(d)** The trajectory (daily cases, 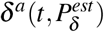).

**Figure 4.**
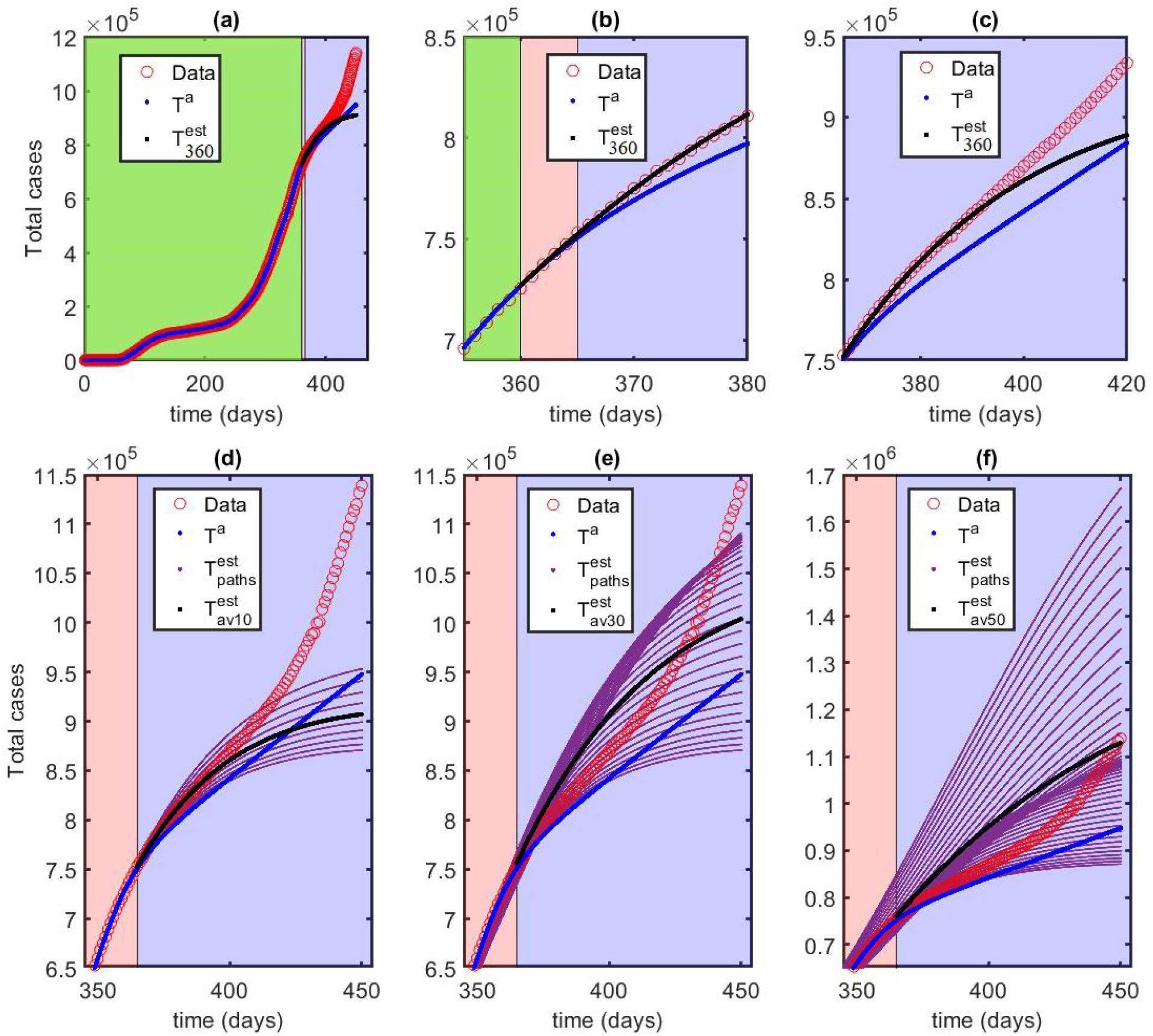
Prediction using previous data: **(a)** Time domain is the first 450 days. *T*^*a*^ represents trial function of total cases; 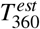 is the prediction starting at the 360th day; red circles are the publicly available data^40^. Time domain has been divided into two parts, testing (green + pink region) and test (light blue region). Narrow pink region is to check the validation of the prediction model. **(b)** Time domain is the first 350 to 380 days of the current pandemic in Canada, focus on the validation (pink) region. **(c)** Time domain is the first 365 to 420 days of the current pandemic in Canada, focus on the test (light blue) region. **(d)** 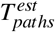 denotes a bundle of 10 path predictions starting points 365th, 364th, …, 356th days of the current pandemic in Canada. 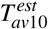 indicates average over 10 path predictions. **(e)** 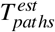 denotes a bundle of 30 path predictions starting points 365th, 364th, …, 336th days of the current pandemic in Canada. 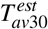 indicates average over 30 path predictions. **(f)** 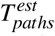 denotes a bundle of 50 path predictions starting points 365th, 364th, …, 316th days of the current pandemic in Canada. 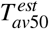 indicates average over 50 path predictions.

**Figure 5.**
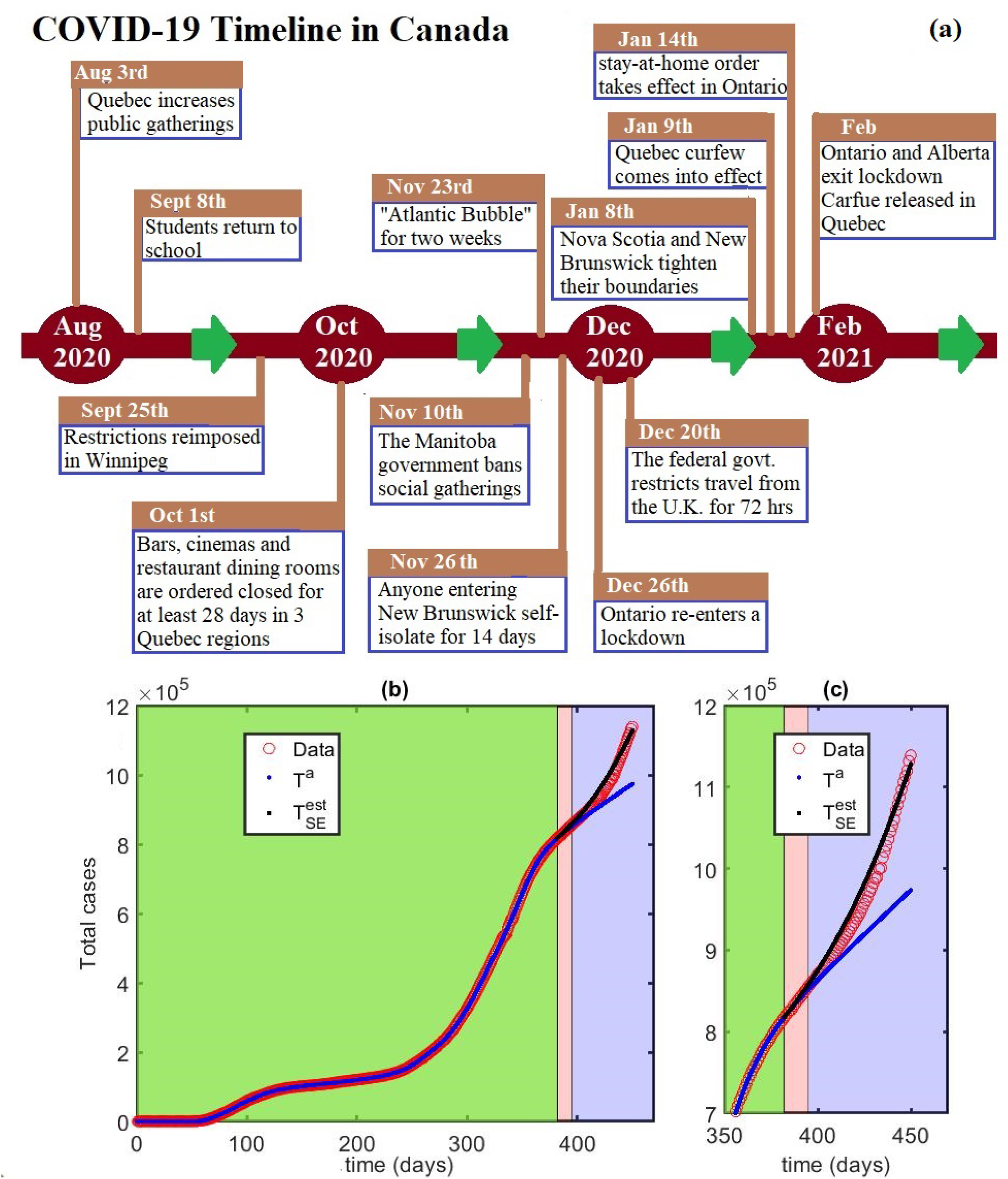
Prediction using previous data: **(a)** COVID-19 timeline in Canada during the period of seven months, August 2020 to February 2021. The image has been generated using Microsoft Paint—Windows 10. **(b)** Time domain is the first 450 days. *T*^*a*^ presents trial function of total cases; 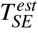 is the prediction using previous social economic condition; red circles are the publicly available data^40^. Time domain has been divided into two parts, training (green + pink region) and prediction (light blue region). Narrow pink region is for check the validation of the prediction model. **(c)** Time scale is the first 350 to 450 days of the current pandemic in Canada, focus on the validation (pink) as well as test (light blue) regions.

The estimated values 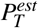 of the parameters, involved in the NN of the trial function of the total cases *T*^*a*^, vary in a wide range of 100 to 300 (Figure 1(a)), and the trial function 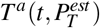 has excellent agreement with the database (Figure 1(b)) during the training period, first 365 days (January 27 2020 to January 25, 2021) of the pandemic in Canada. We analytically compute the first (Figure 1(c)) and second (Figure 1(d)) order derivatives where 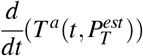 is non negative, as 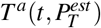 is a monotonic increasing function. However 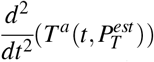 is an oscillating function and can be negative as well as positive.

Using the trial function of total cases and its first and second order derivatives and the second order differential Eq. (5) for total cases, we obtain trial functions 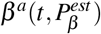 and 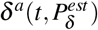 for the model parameters *β* (*t*) and *δ* (*t*) (Figures 2(a) and 2(b)) during the training period of the first 365 days of the current pandemic in Canada. 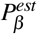 and 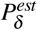 are the estimated values of the parameters *P*_*β*_ and *P*_*δ*_, involved in the trial functions of *β*^*a*^ and *δ*^*a*^, respectively. The range of 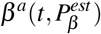 for that period is 0.65 to 0.85. However, over a long interval, first 65th day to 365th day, the value of 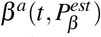 ranges from 0.65 to 0.7. A similar phenomenon occurs for the trial function 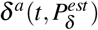; the minimum value of 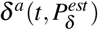 during the training period is 0.4, but over a long interval the range of 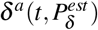 is 0.6 to 0.7. The trial functions and 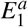 and 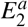 of the error functions *E*_*I*_ and *E*_*S*_ range from 1 to 15 (Figure 2(c)). Using the trial functions 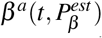 and 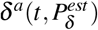, we can calculate the time dependent basic reproduction number *R*_0_(*t*) for the first 365 days of the current pandemic where

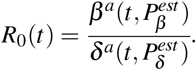

During the interval, first 100th day to 365th day, the value of *R*_0_(*t*) was closed to unity (Figure 2(d)).

From Eq. 4 in the Methods section, it follows that the estimation of the infected population can be expressed by

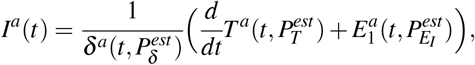

and the number of susceptible individuals can be obtained using the expression

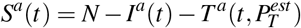

where *N* is the total population size as mentioned in the Methods section. The number of susceptible individuals decreased rapidly after the first 200 days (Figure 3(a)), and the curve of infected individuals shows two peaks (Figure 3(b)), the first peak due to *α*-variant and the second peak due to *β*-variant. The trajectories 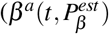, daily cases) (Figure 3(c)) and 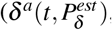, daily cases) (Figure 3(d)) show that after the first 250 days the number of daily cases increase even though the trial functions 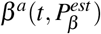 and 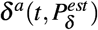 vary in a small range. This phenomenon occurred due to larger number of infected individuals after the first 250 days.

We divide the time domain into three parts (Figure 4(a)); training region, the first 365 days, where we train the trial functions *T*^*a*^(*t*), *β*^*a*^(*t*) and *δ*^*a*^(*t*); validation region, a small interval from the first 360th to 365th day, where we can justify the prediction model; prediction region, an interval from the first 366th day to 450th days. Here, we describe the outcome employing the approach ‘parameters at the fixed point’; a detail is provided in the Methods section. The single path prediction 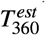 is started at the 360th day of the current pandemic and is extended up to 450th day (Figure 4(a)) along with the trial function of total cases *T*^*a*^. We obtain excellent agreement between 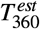 and the database in the validation region (Figure 4(b)). In the initial part of the test domain, the first 365 to 370 days, both curves, 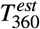 and *T*, have good agreement with the database (Figure 4(c)), and after the initial part, beyond the first 370 days, *T*^*a*^ does not agree with the data. However, we observe an excellent agreement between 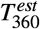 and the data for a wide range of 30 days, the first 366 to 396 days, and at 396th day the difference between *T*^*a*^ and the data is 20,000 which is significantly large. Now, instead of single path prediction, we present the outcome of the multiple path prediction, the 10 path prediction (Figure 4(d)), the 30 path prediction (Figure 4(e)) and the 50 path prediction (Figure 4(f)), along with the trial function *T*^*a*^ and the averages over the multiple paths. Here, the prediction of *M* (= 10, 30, 50) paths represents a bundle, 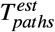, of *M* different predictions on the basis of *M* previous consecutive days, and 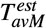 is the overall average of the bundle. 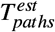 presents a bundle of 10 paths (Figure 4(d)), 30 paths (Figure 4(e)) and 50 paths (Figure 4(f)). The bundle provides an upper and lower bounds of the estimation, and 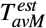 can capture the scenarios, lockdown and reopening, for the last M days. 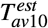 (Figure 4(d)) is the same as the single path prediction 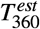 while 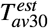 and 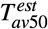 agree with the data of 425th and 450th days, respectively.

The COVID-19 timeline is a combination of several brands of various social economic policies, such as reopening, lockdown, curfew, released curfew, etc. A small part of that timeline (Figure 5(a)), August 2020 to February 2021, shows that due to the increase in public gatherings and the reopening of schools, new COVID-19 cases increased, and to control the situation, the government reimposed various restrictions. After several months in which the situation was under control, the government released those restrictions again. One can observe that pattern multiple times during the current pandemic. It seems that the situation on 382th day of the pandemic was similar to that on 228th day (154 days ago), reopening and releasing restrictions, and we can expect that the social economic policy just after 382th day would be the same as the social economic policy just after 228th days. Therefore, we can assume that the model parameters *β* (*t*) and *δ* (*t*) satisfy the conditions:

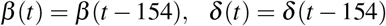

where *t ≥* 382; a detailed is provided in the Methods section. In the calculation we use trial functions 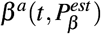 and 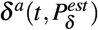 instead of the model parameters *β* (*t*) and *δ* (*t*), respectively. We divide the time domain into three parts; training region, the first 395 days, where we train the trial functions *T*^*a*^(*t*), *β*^*a*^(*t*) and *δ*^*a*^(*t*); validation region, a small interval of the first 382 to 395 days, where we can justify the prediction model 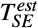; prediction region, an interval of the first 396 days to 450 days. Here, we describe the outcomes 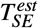 employing the approach ‘parameters map previous social economic situation’; a detail is provided in the methods section. We obtain an excellent agreement between 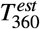 and the database in the validation region (Figure 5(b)). We observe outstanding agreement between 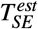 and the database in the validation region (Figure 5(c)). We and the data. observe an outstanding agreement (Figure 5(c)) between 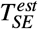 and the data for a wide range of 54 days, the first 396 to 450 days, and at 450th day there is a large discrepancy between *T*^*a*^ and the data.

## Discussion

The function 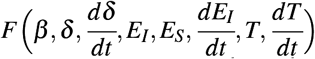, the third term of the prediction model, is equal to 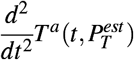, which can be either positive or negative (Figure 1(d)), indicates how social economic policy affects the COVID-19 cases. Various restrictions such as lockdown, curfew decrease the new cases; contrastingly, reopening, releasing restrictions increase the new cases. Social economic policy plays the role of maintaining a balance between COVID-19 cases and socioeconomic constraints, such as not a long lockdown, not closing businesses for a long time, and not letting daily cases are skyrocketing.

Initially, there was no social economic constraint due to COVID-19, all susceptible individuals did not know about the disease because of that the value of 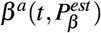 was high. At the beginning, COVID-19 test did not progressed, i.e., it took time to provide a large scale covid test, and a part of the public did not know about it. The symptoms of COVID-19 are similar to those of the common flu that confused the population. As a consequence, the value of the function 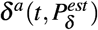 was significantly low. Due to the reasons mentioned above, initially, the basic reproduction number *R*_0_(*t*) was higher compared to the time interval, first 100 to 365 days, where *R*_0_(*t*) was closed to unity. For a particular time *t*, the pair 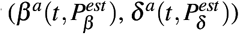 corresponds to a certain socioeconomic condition; in other word, the trial functions of the model parameters *β* (*t*) and *δ* (*t*) can capture the social economic policy.

The ‘prediction using previous data’ 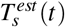 is based on the socioeconomic conditions of the starting time *s* and forecasting for the time domain *t > s*. The prediction shows the expected outcomes if the socioeconomic conditions remain the same as at the start time; in single path prediction, we consider *β*^*a*^(*t*) = *β*^*a*^(*s*) and *δ*^*a*^(*t*) = *δ*^*a*^(*s*) for *t > s*. Since *β*^*a*^(*t*) and *δ*^*a*^(*t*) are slowly varying functions, the single path prediction 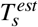 estimates perfectly over a short period. The multiple path prediction is a collection of single path predictions and can provide the range of the highest and lowest possibilities, depending on the situations of the start times of the single path predictions. The average over multiple path (*M* paths) prediction 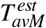 takes into account the general socioeconomic conditions during the period of *M* paths. In the present calculation *M* path prediction is the bundle of *M* single path predictions with starting points *M* consecutive days because we use a daily basis public database.

The lockdown/socioeconomic restriction and reopening/release of the socioeconomic restriction happened and are happening one after another throughout the current pandemic. The repetition of the events is the principle concept to use the previous situation to predict the continuing scenario. Naturally, the proceeding situation will not be exactly the same as the previous situation. However, the earlier situation can help to predict the growing situation.

## Conclusion

In this article, we proposed a hybrid approach to forecast the COVID-19 cases using previous database of total cases. For this purpose, we derived a second order nonlinear differential equation, involved the time dependent model parameters, for the total cases; this differential equation is the foundation of the present study. The trial function of total cases has been generated using a single layer NN and publicly available database. The first and second order derivatives of the total cases have been evaluated analytically using that trial function. Using inverse problem technique on the second order nonlinear differential equation, we obtained trial functions for model parameters. We used Taylor series expansions to derive four kinds of prediction: single path, multiple path, average over multiple path and using previous socioeconomic phenomena. To assess the prediction models, we divided the time domain into two parts, training and test. We estimated the parameters of NNs, i.e., we train the error functions during the training period. In the test domain, the prediction models show excellent agreement with the data. The present work is free to solve any system of differential equations because of that the proposed hybrid method is computationally simple. In future work, we will consider COVID-19 dynamics for two neighbouring states with moving population. One can study the recovery and death toll rates using the second order differential equations mentioned in the Methods section. The approach used in this work could be applied in the study of other infectious disease transmission dynamics as well.

## Methods

In this section, we introduce a compartment based infectious disease model, extending the standard SIR model, including a total of five partitions, Susceptible, Infected, Confirmed cases, Recovered and Deaths (SICRD). The model is constructed as a set of coupled ordinary differential equations involving several time dependent parameters. The entire methodology is a combination of four parts:

- **Step 1 :** We generate a second order nonlinear differential equation for the total cases (confirmed cases + recovered + deaths) that involves two time dependent model parameters.
- **Step 2 :** We decompose the time domain into training and test intervals, and develope a trial function for the total number of cases using NN. The parameters involved in NN are estimated to minimize the error function over the training domain. Thereafter, we calculate the derivatives of that trial function analytically.
- **Step 3 :** Solving the inverse problem for the differential equation, from Step 1, using the trial function and its derivatives, from Step 2, we obtain trial functions for the time dependent model parameters, from Step 1.
- **Step 4 :** Finally, we develop several prediction models for total cases using Taylor’s expansion. The variety of the prediction models is based on the choice of the time dependent model parameters.

### The Model

Modeling the spread of epidemic is an essential tool for projecting its outcome. By estimating important epidemiological parameters using the available database, we can make forecasts of different intervention scenarios. In the context of compartment based model, where the population of a region is distributed into several groups, such as susceptible, infected, confirmed cases, etc., is a simple but useful tool to demonstrate the panorama of an epidemic.

The schematic diagram of the model is presented in Figure 6(a) with several compartments and time dependent model parameters. The following are the underlying principles of the present model.

**Figure 6.**
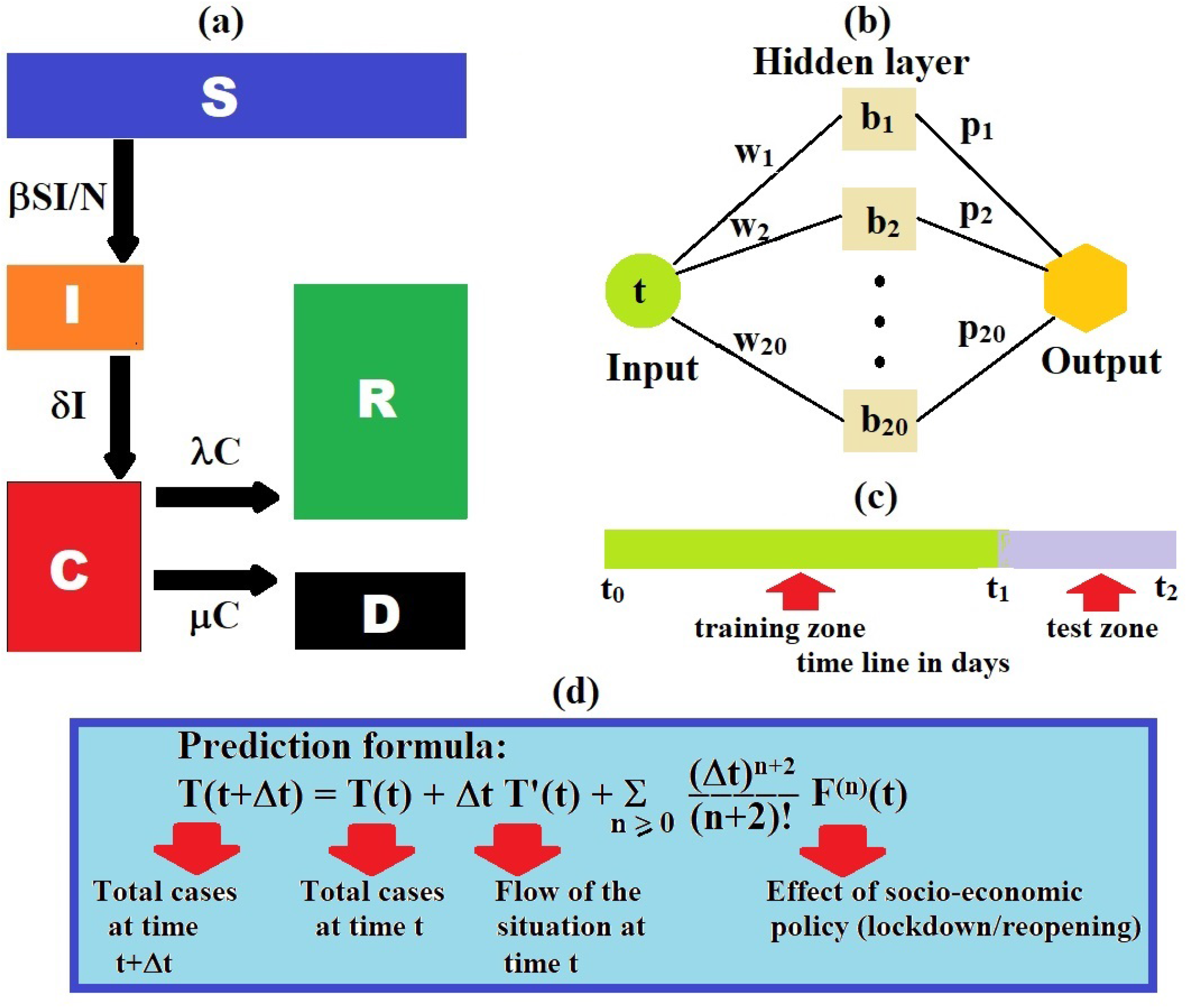
Model and methodology: **(a)**Schematic diagram of the compartmental based epidemic model. **(b)** Structure of the single hidden layer NN **(c)** Time domain has been divided into two parts, training and test. **(d)** The proposed prediction model, outcome of the present study.

- The total population is constant (neglecting the migrations, births and unrelated deaths) and initially every individual is assumed susceptible to contract the disease.
- The disease is spread through the direct (face-to-face meeting) or indirect (through air current, common used or delivery items like door handles, grocery products) contact of susceptible individuals with the infected individuals.
- The quarantined area or the compartment for corona cases contains only members of the infected population who are tested corona-positive.

- The virus always kills some percent of the people it infects; the survivors percent represents the recovered group. Based on the above principles, we consider several compartments:
- Susceptible (*S*): the group of individuals who can be infected.
- Infected (*I*): the group of people who are spreading the contiguous disease.
- Confirmed cases (*C*): the group of individuals who tested corona-positive.
- Recovered (*R*): the group of recovered individuals who tested corona-positive.
- Deaths (*D*): the group of deaths individuals who tested corona-positive.
- Total cases (*T*): the group of individuals who tested corona-positive (Confimed cases + Recovered + Deaths).

In the model, we assume that there is no overlap between these two compartments, infected(*I*) and total cases (*T*). In other words, tested corona-positive individuals are assumed to be unable to substantially spread the disease due to isolation and are immune to re-infection after recovery^41^. The time-dependent model is the following set of coupled delay differential equations:

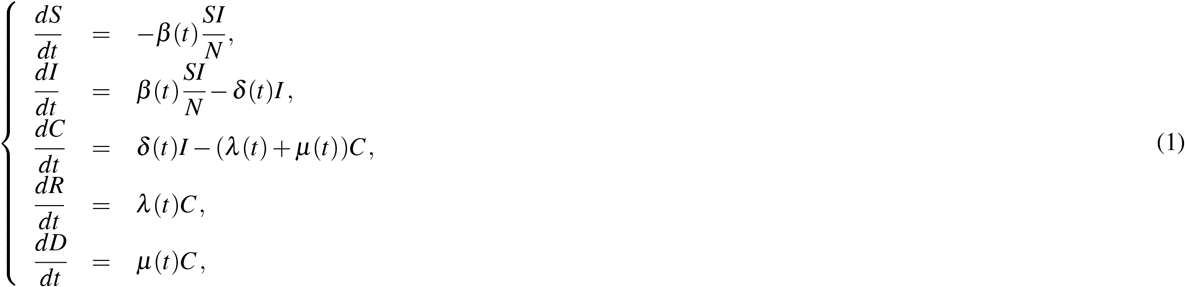

where *β* (*t*), *δ* (*t*), *λ* (*t*) and *µ*(*t*) are real positive parameters, respectively, modeling the rate of infection, the rate of tested corona-positive, the rate of recovery and the rate of death toll. It follows from (1), that for any time *t*

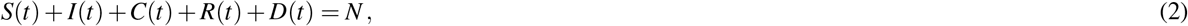

where *N* (constant) is the total population size. Using the fact, total cases *T* = *C* + *R* + *D* the system of equations (1) can be written as

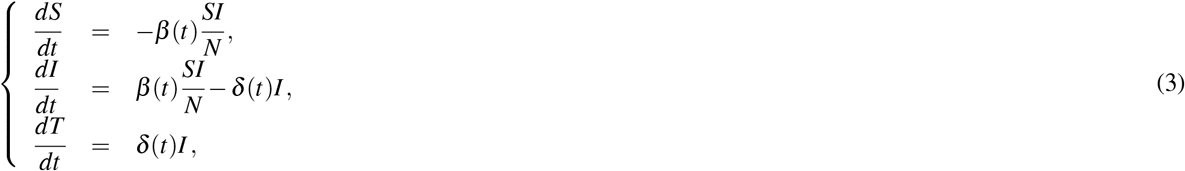

It follows from third equation of (3)

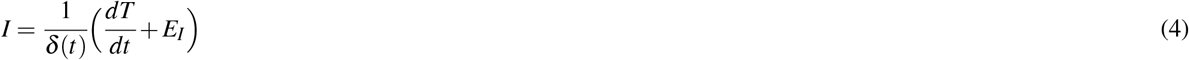

where *E*_*I*_ is the model correction associated with *I*. The cause of model correction is the inherent simplifications and approximations that are made in postulating a mathematical model of a physical phenomenon, as even high-fidelity models simply approximate reality. These simplifications may arise from lack of knowledge about the system and/or computational constraints. Eliminating *S* from (3) and using (4), we obtain a second order nonlinear ordinary differential equation for *T* involving the time dependent parameters *β* (*t*) and *δ* (*t*) as

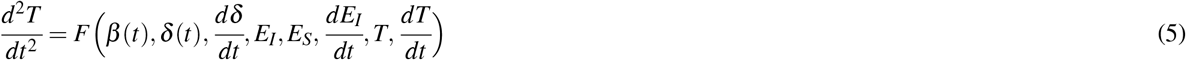

where

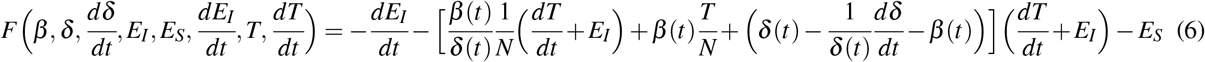

and *E*_*S*_ is the model correction associated with *S*.

It follows from the last three equations of (1) and third equation of (3):

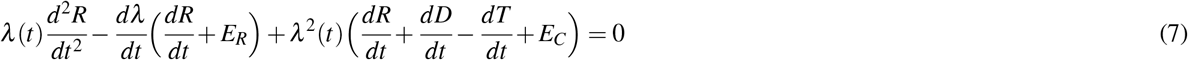

and

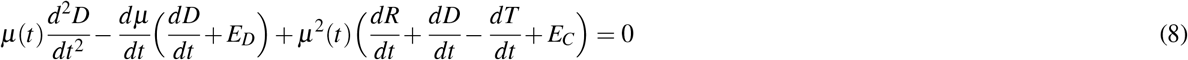

where *E*_*R*_, *E*_*D*_ and *E*_*C*_ are the model corrections. The above two Eqs. (7) and (8) can be used to calculate the recovery rate *λ* (*t*) and the rate of death toll *µ*(*t*). However, the present work is devoted for forecasting the COVID-19 cases so we merely focus on (5), the second order differential equation for total cases.

### The trial function

In this section, we construct a trial function for *T* (*t*), and the parameters of this function are estimated using publicly available database. The trial function for *T* (*t*), satisfied the Eq. (5), may be written in the following form:

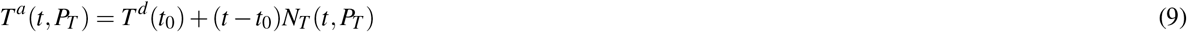

above function has two parts, the first part satisfies the initial condition (at *t* = *t*_0_) and contains no adjustable parameters, the second part involves a NN *N*_*T*_ (*t, P*_*T*_) (Figure 6(b)) containing adjustable parameters *P*_*T*_. Here, *T*^*d*^(*t*_0_) indicates the observed data, total cases at time *t*_0_, obtained from the publicly available database. The NN *N*_*T*_ (*t, P*_*T*_) that takes a single input value of time t is defined as follows:

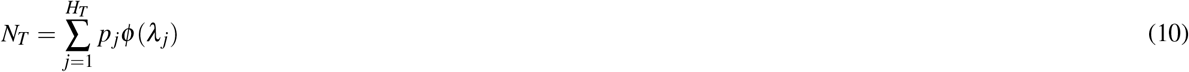

where the sigmoid activation function

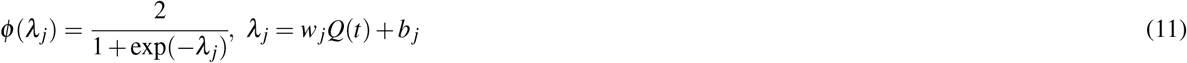

and

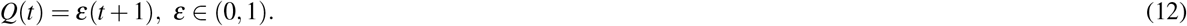

The NN *N*_*T*_ (*t, P*_*T*_) has one hidden layer with *H*_*T*_ activation functions and a linear activation function in output unit. Here, *w*_*j*_ is a weight parameter from input layer to the *j*th hidden layer, *b* _*j*_ is a bias for the *j*th hidden layer, *p* _*j*_ is a weight parameter from the *j*th hidden layer to the output layer. Here, we choose the sigmoid activation function, presented in Eq. (11), because *T* (*t*) is a monotonic increasing function.

The NN should be trained using the publicly available database. Therefore, the time domain is divided into two parts which are training, *t*_0_ *≤ t ≤ t*_1_, and test, *t*_1_ *< t ≤ t*_2_ (Figure 6(c)). Using the publicly available data 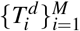 corresponding the M discrete points in time domain [*t*_0_, *t*_1_] with step length *h*, the NN is trained by finding the optimal values of the weight and bias parameters (*p*_*j*_, *w*_*j*_ and *b* _*j*_) which provide the minimum value of the root mean square error function. The error function *ℰ* _*T*_ (*P*_*T*_) has the following form:

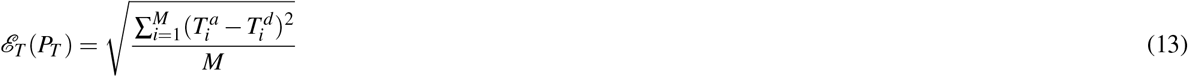

where 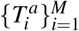 are the values of the trial function *T*^*a*^(*t, P*_*T*_) corresponding the M discrete points in time domain [*t*_0_, *t*_1_] with step. We can express 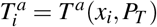 where *x*_*i*_= *t*_0_+ (*i−* 1)*h* for *i* = 1, 2, …, *M* and *x*_1_ = *t*_0_, *x*_*M*_ = *t*_1_. The sufficient small values of *ϵ* _*T*_ (*P*_*T*_) indicates the estimated values of *P*_*T*_, denoted as 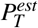.

### The inverse problem

In this section, we derive the trial functions for *β* (*t*) and *δ* (*t*), and the parameters of these trial functions are estimated using inverse problem techniques. After computing the trial function 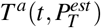, we calculate the first and second order derivative of 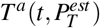 analytically for the time domain [*t*_*0*_, *t*_*1*_] The trial function for *β* (*t*) defined in the time domain [*t*_*0*_, *t*_*1*_], involved in the Eq. (5), may be written in the following form:

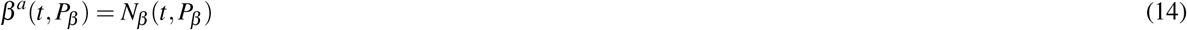

above NN *N*_*β*_ (*t, P*_*β*_) contains adjustable parameters *P*_*β*_. The NN *N*_*β*_ (*t, P*_*β*_) that takes a single input value of time t is defined as follows:

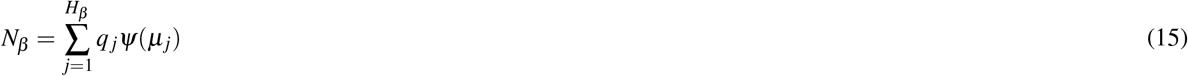

where the hyperbolic activation function

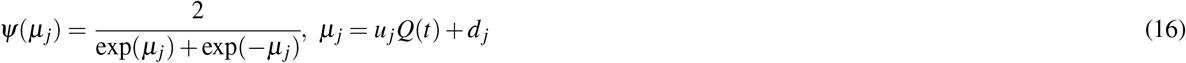

and

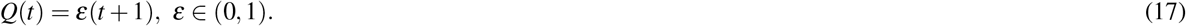

The NN *N*_*β*_ (*t, P*_*β*_) has one hidden layer with *H*_*β*_ activation functions and a linear activation function in output unit. Here, *u* _*j*_ is a weight parameter from input layer to the *j*th hidden layer, *d* _*j*_ is a bias for the *j*th hidden layer, *q* _*j*_ is a weight parameter from the *j*th hidden layer to the output layer. Similar type of NNs can be used as the trial functions for *δ* (*t*), *E*_*I*_ and *E*_*S*_, involved in the Eq. (5). The error function associated with the inverse problem that must minimize, has the following form:

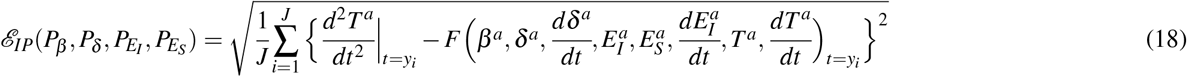

where 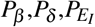 and 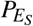 are the parameter sets associate with the trial functions 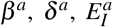 and 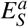, respectively. The discrete points *y*_*i*_ for *i* = 1, 2,, *J* can be expressed as *y*_*i*_ = *t*_0_ + (*i* − 1)Δ*t* and *y*_1_ = *t*_0_, *y*_*J*_ = *t*_1_. Here, error function 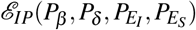 for *J* equally spaced points with step length Δ*t* inside the interval [*t*_0_, *t*_1_] is trained. The sufficient small values of 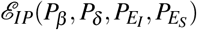 indicates the estimated values of 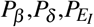 and 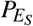 denoted by 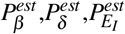 and 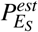, respectively.

### Prediction

In this section, we develop prediction models for the total number of cases using Taylor series. Suppose that total confirmed cases at time *t*_1_ i.e., *T* (*t*_1_) is given, and we want to predict *T* (*t*_1_ + Δ*t*) i.e., total confirmed cases at time *t*_1_ + Δ*t*. In this context, we can use Taylor series method for *T* (*t*_1_ + Δ*t*).

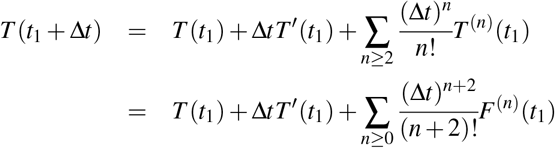

where *T* ^*′*^(*t*_1_) and *T* ^(*n*)^(*t*_1_) are the first and n-th order derivatives. Here, we use the fact *T* ^(*n*+2)^ = *F*^(*n*)^, obtained from the Eq. (5). The right hand side of the final expression for *T* (*t*_1_ + Δ*t*), presented in Eq. (19), has three terms (Figure 6(d)).

- *T* (*t*_1_) : Total cases at time *t*_1_
- Δ*tT*^*′*^(*t*_1_) : Effect of the situation at time *t*_1_ on time *t*_1_ + Δ*t*
- 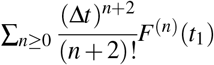: Effect of socio-economic policy, i.e., lockdown or reopening; the pair (*β* (*t*_1_), *δ* (*t*_1_)) represents a particular event/situation such as lockdown or reopening.

For simplicity we neglect all the derivatives of the function *F (t*_*1*_) and replacing 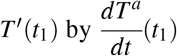, and obtain an estimated value for *T* (*t*_1_ + Δ*t*), denoted by *T* ^*est*^(*t*_1_ + Δ*t*), as follows

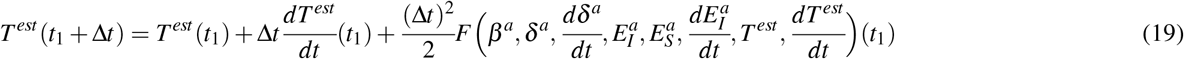

where at time *t*_1_ *T* ^*est*^(*t*_1_) = *T* (*t*_1_) and 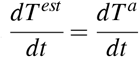. Again using Taylor series expansion, we can express

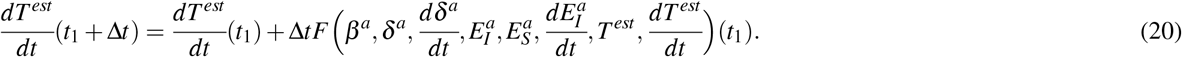

Using the recurrence relations, Eqs. (19) and (20), we can predict the total corona positive cases for the time domain *t≥ t*_1_ + Δ*t*. According to the choice of the trial functions *β*^*a*^ and *δ*^*a*^, we can derive two types of prediction model.

- To consider the fixed values of *β*^*a*^ and *δ*^*a*^, i.e., for the prediction domain *t ≥ t*_1_ + Δ*t*, and

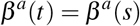

and

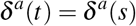

where *s* belongs to the training domain.
- To consider the previous time evolution for *β*^*a*^ and *δ*^*a*^, i.e., for the prediction domain *t ≥ t*_1_ + Δ*t*, and

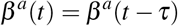

and

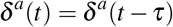

where *t − τ* belongs to the training domain. Here, *β*^*a*^ and *δ*^*a*^ correspond to the previous social economic situation.

### Parameters at the fixed point

In this section, we discuss the prediction models for fixed values of the trial functions *β*^*a*^ and *δ*^*a*^. Suppose the time domains for training and test are [*t*_0_, *t*_1_] and (*t*_1_, *t*_2_] (Figure 6(c)), respectively, and we use the Eqs. (19) and (20) and a starting point to estimate the values of *T*, defined as 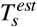 where the starting time *s* belongs to the interval (*t*_0_, *t*_1_], for the second domain. Here, previous data means that we consider the same social economic condition as the starting time *s* through out the calculation. As a consequence in Eqs. (19) and (20), the functions 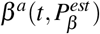 and 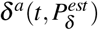 are replaced by 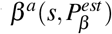 and 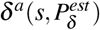 respectively, for *t*_0_ *< s ≤ t*_1_. We can obtain an estimated value for 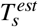 as follows

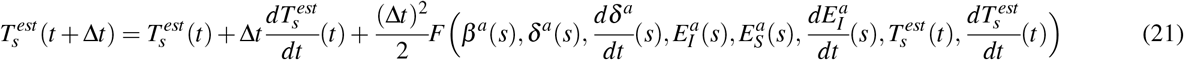

where at the starting time 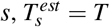 and 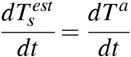 Again using Taylor series expansion, we can express

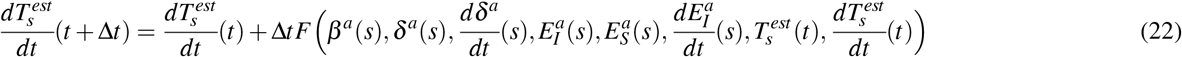

where *s ≤ t ≤ t*_2_. Here, we neglect the time evolution of the functions 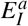 and 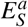.

The starting points for the estimation can be *t*_1_ *−k*Δ*t, k ≥* 0, and the prediction starting through the point *t*_1_ *−k*Δ*t* can be defined as *k*-th path and denoted by 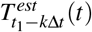 for *t*_1_ *< t ≤ t*_2_. The first *K* paths are as follows:

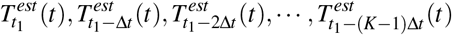

The bundle of the first *K* paths can be denoted by 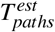 Taking average over first *K* paths, we obtain estimation of total cases *T* over *K* paths as follows:

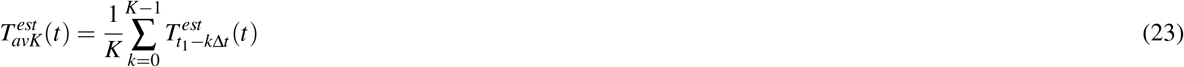

where *t*_1_ *< t ≤ t*_2_.

### Parameters map previous social economic situation

In this section, we demonstrate the prediction model when the trial functions *β*^*a*^ and *δ*^*a*^ correspond to the previous Social Economic (SE) situation. Suppose the time domains for training and test are [*t*_0_, *t*_1_] and [*t*_1_ + Δ*t, t*_2_] (Figure 6(c)), respectively, and we use the Eqs. (19) and (20) and a previous SE situation to estimate the values of *T*, defined as 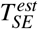 for the second domain. Here, we assume that the social economic condition of the period [*t*_1_ + Δ*t, t*_2_] is similar with the social economic condition of *τ* days ago. As a consequence in Eqs. (19) and (20), the functions 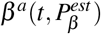 and 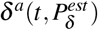 are replaced by *β*^*a*^(*t −τ*) and*δ*^*a*^(*t −τ*), respectively, for *t*_1_ + Δ*t ≤ t ≤ t*_2_ and *t*_0_ *≤ t −τ ≤ t*_1_. We can obtain an estimated value for 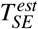,as follows

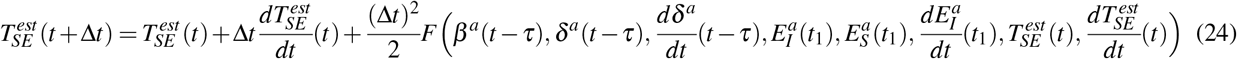

where at time 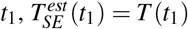 and 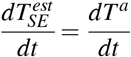. Again using Taylor series expansion, we can express

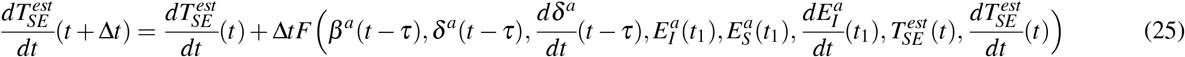

where *t*_1_ *< t ≤ t*_2_. Here, we neglect the time evolution of the functions 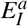 and 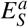.

### Numerical experiment

In this section, we propose a detailed description of the computational procedure for the proposed approach. To justify the present method i.e., training, validation and test purpose we consider Canadian COVID-19 database where the training period is the first 365 days of the current pandemic, from January 27, 2020 to January 25, 2021. The simulation is conducted with matlab, and the in build function **fminunc** is used to minimize the error functions. The initial values of the parameters are selected randomly. To calculate the trial function *T*^*a*^ for total confirmed cases, we consider *H*_*T*_ = 20 i.e., 20 sigmoid units in the hidden layer, *ε* = 0.1, and *h* = 1 due to daily basis publicly available COVID-19 database. We obtain an estimated value 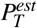 of *P*_*T*_ for corresponding the value of the error function 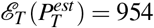. To calculate the time dependent functions 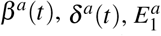 and 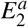 efined in the training period, we consider 30 hyperbolic secont units in the hidden layer, *ε* = 0.1 and the step length Δ*t* = 0.1. We obtain an estimated value 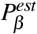 of 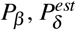 of 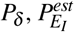 of 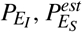 of 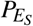 for corresponding the value of the error function 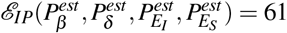. For the prediction of total confirmed cases using Taylor series expansion method, we consider the step length Δ*t* = 0.1.

## Data Availability

The data used to estimate the model parameters are publicly available and are available with the code.

https://resources-covid19canada.hub.arcgis.com/datasets/case-accumulation/data.

## Code availability

All code is available in the GitHub repository for the project at https://github.com/SPAUL2021/COVID19PREDICTION

## Acknowledgements

This research is supported by the Natural Sciences and Engineering Research Council of Canada (NSERC).

## Author contributions statement

S.P. has derived the second order differential equations, the prediction models and developed the matlab codes, has analyzed the calculated results, and has prepared all figures. S.P. and E.L. have drafted the original article. Both authors have contributed to the editing of the article. Both authors have read and approved the final article.

## Additional information

Correspondence and requests for materials should be addressed to S.P.

